# Fragmented and Underutilized: Exploring Role Variation and Ambiguity for Research Project Managers in Collaborative Team Science

**DOI:** 10.64898/2026.10.26.26366534

**Authors:** Shayla A Vradenburgh, Betsy Rolland

**Affiliations:** Michigan Institute for Clinical and Health Research, University of Michigan, Ann Arbor, MI, United States

**Keywords:** Research project management_1_, project management_2_, clinical and translational research_3_, academic research_4_, team science_5_

## Abstract

**Introduction:** Clinical and translational research (CTR) increasingly relies on collaborative and interdisciplinary research teams to translate discoveries into practical solutions. Research project managers (RPMs) often support CTR projects to ensure continual progress toward research goals, yet there is still ambiguity around what research project management is and the role RPMs play on research teams. Our study aimed to address this gap in knowledge by understanding how RPMs (1) define project management in an academic research context and (2) align on the key job responsibilities of RPMs.

**Methods:** Survey responses from 85 academic RPMs and 14 semi-structured follow-up interviews were analyzed to characterize their perspectives and experiences.

**Results:** Findings revealed four themes: (1) RPMs were in alignment with each other and the Project Management Institute on the definition of project management while noting distinctions between academic and industry contexts, (2) RPMs reported diverse job responsibilities despite having similar job titles, (3) RPMs struggled with role ambiguity stemming from variability in key job responsibilities, and (4) RPMs identified areas of support to improve research project management.

**Discussion:** These findings contribute to a greater understanding of project management in academic research contexts and highlight the variable utilization of RPMs on collaborative teams. Well managed projects rarely happen naturally, but can have a large impact on research environments, team member wellbeing, and overall research productivity. By characterizing how RPMs define and enact their role, this study provides a foundation for developing resources and avenues of support that can ultimately increase the efficiency of translating research discoveries from bench to bedside.

## 1 Introduction

A primary component of clinical and translational research (CTR) in academia is conducting research projects, yet there is minimal guidance or training in how to effectively move a project from start to finish. The process of getting research knowledge from the bench to the bedside is long, with some studies reporting only ∼10% of scientific discoveries are translated into practice within 20 years. The complete lifecycle of translating research findings often requires sufficient progress and conclusions on multiple different projects, where poor management of any individual project in that pipeline can lead to a compounding delay in translation of promising health discoveries (Drolet & Lorenzi, 2011; Contopoulos-Ioannidis et al., 2003).

The field of project management (PM) is dedicated to helping improve the efficient and successful completion of projects. The Project Management Institute (PMI) defines project management as “the application of knowledge, skills, tools, and techniques to project activities to meet project requirements” (Project Management Institute, n.d.-b) and has the stated organizational purpose to “maximize project success to elevate our world.”(Project Management Institute, n.d.-a). One way the PMI contributes to this mission is through the creation and maintenance of the Project Management Body of Knowledge (PMBOK). Initially published in 1986 with the latest edition being released in 2025, the PMBOK Guide serves as an internationally recognized publication that standardizes PM principles and best practices (Project Management Institute, 2026; Webster, 1994). While the PMBOK Guide serves as a valuable resource and creates some standardization in the field of project management, most recommendations are better suited for industries outside of academia like construction and software development (Powers & Kerr, 2009; Svejvig et al., 2025).

While inspiration and foundational PM knowledge can be drawn from broader organizations like the PMI, some of the differences between academia and industry make conventional advice less effective for managing academic research projects. Some of the qualities of academic biomedical research that make its environment distinct from industry settings include less concrete objectives and deliverables, more uncertainty and need for flexibility, and diffuse authority and accountability (Beukers, 2011; Powers & Kerr, 2009). Gaining a better understanding of how project management is defined and applied in a research setting can inform efforts to better support and enhance collaborative projects (Pacheco Junior et al., 2020; Payne et al., 2011).

As the complexity of collaboration in research projects increases, research groups and funding agencies are starting to acknowledge the value project management can provide for CTR teams. For example, grant proposals for programs like the National Science Foundation’s (NSF) Science and Technology Centers: Integrative Partnerships must include a management plan. Unlike the leadership plans required by some NIH grants with multiple PD/PIs, management plans typically encompass the broader team and involve identifying key members of the management team and detailing how project work will be operationalized (U.S. National Science Foundation, 2024). Grant requirements, along with an increased appreciation for the value project management can provide research projects, has led to many research groups hiring dedicated project managers. While funding agencies and research groups may see the value in hiring a professional to help with managing a research project, there is still little standardization around what research project management is and the core responsibilities a research project manager (RPM) should have.

In this manuscript, we address the gap in knowledge that exists around what research project management is and what RPMs do by answering the following questions from the perspective of professionals currently working as RPMs.

1. How are RPMs’ definition of project management in alignment with one another and the definition provided by the PMI, and how does project management in academic research differ from other industries?
2. How similar are job responsibilities described by RPMs and what challenges arise from the variability in job expectations?
3. What additional support do RPMs want to succeed in their roles and how can we enhance the field of research project management?

## 2 Materials & Methods

### 2.1 Recruitment

This study used a broad recruitment campaign to recruit survey and interview participants, including posts and directed outreach via LinkedIn, email outreach to administrators and researchers involved with various Clinical and Translational Science Award (CTSA) Hubs, groups like the NIH Science of Team Science (SciTS) and National Organization for Research Development Professionals (NORDP) listservs, and reaching out to personal networks of researchers and administrators to participate.

### 2.2 Data Collection

Eligibility criteria for the survey administered through Qualtrics included having the title of project manager (or equivalent) for a funded scientific research project. Based on confirmation of meeting the eligibility criteria and self-selection of their current job title as “research project manager (or equivalent title),” participants were sent a link to the survey (Supplementary Material).

Following completion of the survey, respondents had the opportunity to participate in a virtual, 1 hour, semi-structured follow-up interview to further discuss the challenges they faced around project management, the tools they used to manage their research projects, and resources they would like to have adapted for their research environment. Interviews were conducted via Zoom, recorded, and transcribed. Transcripts were anonymized and assigned pseudonyms to protect participant’s identities. This study was deemed exempt by the University of Michigan’s Institutional Review Board, HUM00255826.

### 2.3 Data Analysis

When analyzing Likert scale survey data, the average score and standard deviation for all respondents are reported.

Interview responses were coded using thematic analysis in NVivo 15. Responses could be coded to more than one theme. Specifically when coding PM responsibilities, the broader themes for management-related tasks were created by referencing the performance domains found in the PMI’s A Guide to the Project Management Body of Knowledge (PMBOK Guide, Eighth Edition). Based on interview responses and some of the themes referenced in previous studies (Venkatesh & Rolland, 2025), we made a few modifications. The broader category of the resource performance domain was separated into a more specific code of managing “physical resources”. We also combined the resource and stakeholder performance domains to create a new code of managing “team members”. Lastly, we separated out the “information management” code from the broader governance performance domain in alignment with our ongoing research (Venkatesh & Rolland, 2025).

Only survey and interview data relevant to the stated research questions associated with this manuscript are analyzed and reported here. Additional findings from this data will be explored in future publications.

### 2.4 Participants

A total of 85 respondents self-identified as academic RPMs or equivalent roles (people who manage project planning, coordination, and execution). Sixty-seven (78.8%) participants identified as female, 77 (90.6%) identified as Not Hispanic or Latino, 70 (82.4%) identified as White (Table 1), and a variety of research types were represented among participants (Table 2). Of the 85 survey respondents, 14 agreed to participate in a follow-up interview.

**Table 1:** Survey participant demographics.

| Demographic Category | # (%) |
| --- | --- |
| **Gender** |  |
| Female | 67 (78.8%) |
| Male | 17 (20%) |
| Prefer not to answer | 1 (1.2%) |
| **Ethnicity** |  |
| Hispanic or Latino | 5 (5.9%) |
| Not Hispanic or Latino | 77 (90.6%) |
| Prefer not to answer | 3 (3.5%) |
| **Race** |  |
| Asian | 6 (7.1%) |
| Black or African American | 5 (5.9%) |
| White | 70 (82.4%) |
| White, Black or African American | 1 (1.2%) |
| Prefer not to answer | 3 (3.5%) |

**Table 2:** Survey participant research types.

| Research Type | # (%) |
| --- | --- |
| Basic | 8 (9.4%) |
| Basic, Clinical | 2 (2.4%) |
| Basic, Clinical, Community-engaged | 1 (1.2%) |
| Basic, Clinical, Translational | 4 (4.7%) |
| Basic, Clinical, Translational, Community-engaged | 6 (7.1%) |
| Basic, Clinical, Translational, Community-engaged, Other | 1 (1.2%) |
| Basic, Community-engaged | 3 (3.5%) |
| Basic, Translational | 6 (7.1%) |
| Basic, Translational, Community-engaged | 5 (5.9%) |
| Clinical | 10 (11.8%) |
| Clinical, Community-engaged | 4 (4.7%) |
| Clinical, Translational | 6 (7.1%) |
| Clinical, Translational, Community-engaged | 5 (5.9%) |
| Community-engaged | 3 (3.5%) |
| Community-engaged, Other | 1 (1.2%) |
| Other | 5 (5.9%) |
| Translational | 7 (8.2%) |
| Translational, Community-engaged | 8 (9.4%) |

Thirty-six (42.4%) survey respondents have had their current job for 3-5 years (Figure 1A) and 43 (50.6%) shared that they have received formal PM training (Figure 1B). Interview participants identified some of the ways they have received PM training which include institutional training/coursework, online training, trial-and-error methods or self-taught, and training through formal project management organizations like the PMI (Figure 1C).

**Figure 1:**
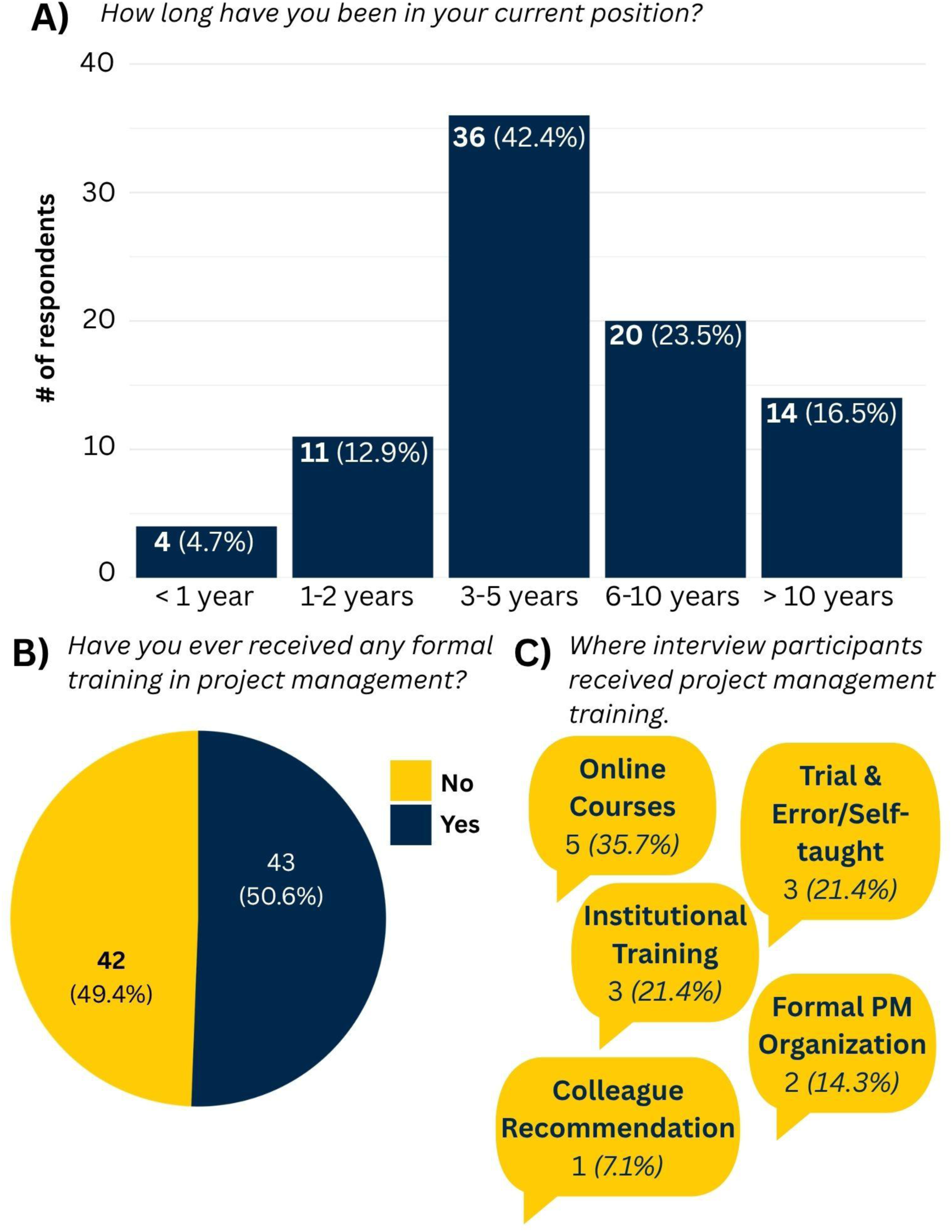
A) Distribution of survey respondents by time in their current position. **B)** Proportion of survey respondents who reported receiving formal project management training. **C)** Sources of project management training reported by interview participants. For all panels, the first number indicates the number of respondents, with percentages in parentheses representing the proportion of all respondents.

## 3 Results

### 3.1 RPMs were aligned on their definition of project management while highlighting how project management is distinct in academic versus industry settings

Survey respondents showed alignment with the PMI definition of project management, with interview participants sharing additional insights. Survey participants reported general alignment with the PMI definition of project management with a mean score of 4.07 (standard deviation 0.92). Interview participants further defined project management, with responses falling into four themes: defining, managing, navigating, and optimizing (Table 3).

**Table 3:** How would you define project management?

| Code | Illustrative Quote | # of participants (%) |
| --- | --- | --- |
| Defining | “Ideally it’s helping project teams scope and define the project, define deliverables, and timeline objectives to get [a defined project and deliverables]” - *Nora* | 4 (28.6%) |
| Managing | “Understanding and overseeing and being responsible for making sure that all the various requirements of a specific project happen.” - *Bethany* | 12 (85.7%) |
| Navigating | “[R]esearch project management is about navigating teams through ambiguity and providing structure. And making it so that... changes and ambiguity are not causes for chaos, or alarms, or obstacles, but are natural... parts of the process that a team navigates with the support of a project manager.” - *Nicole* | 3 (21.4%) |
| Optimizing | “What I think of when I think of project management is organizing and streamlining workflows.” - *Lacy* | 2 (14.3%) |

Four (28.6%) interview participants shared descriptions of project management that included defining project objectives, tasks, and performance areas including cost, scope, timelines, and resources. One interview participant summed up their definition of project management as “helping project teams scope and define the project, define deliverables, and timeline objectives to get [a defined project and deliverables]” (Nora). Twelve (85.7%) interview participants shared responses that align with management of projects. Their responses included oversight of the full project lifecycle, assuring goals are completed, tracking task completion and project progress, managing project and grant activities, coordinating people, and managing project cost, scope, timelines, and resources. Three (21.4%) interview participants shared definitions of project management that included navigating teams through ambiguous and changing projects as well as risks. This PM definition was well summarized by one interview participant who said, “[R]esearch project management is about navigating teams through ambiguity and providing structure. And making it so that… changes and ambiguity are not causes for chaos, or alarms, or obstacles, but are natural… parts of the process that a team navigates with the support of a project manager (Nicole)”. Two (14.3%) interview participants described project management in terms connected with optimizing or improving workflows.

Five (35.7%) interview participants noted the ways in which project management in academic research environments differs from project management in other industries (Table 4):

1. Project management in academic research prioritizes discovery and innovation over product creation, which is often the primary focus of other industries. “I feel like so many of the PMI-specific resources are so tailored to software, construction, [where] you are creating a specific thing,” shared Kelli. As many research projects don’t have the ultimate goal of creating a specific product, it can be challenging to fit industry standard resources into the academic research environment.
2. Academic research requires more flexibility in PM practices as project elements can often change. Kevin reflected on this difference by sharing about their previous work experience managing projects in the software industry. “I worked previously in software development, so I’m very much focused on, like, let’s create a timeline and then let’s meet these deadlines and deliverables. And it seems like research requires a certain amount of flexibility for tweaking how you’re going to meet the aims of your grant… It became more frustrating to try and keep this project timeline map that was historically very important for my work.” The tools and strategies that previously were helpful in another industry became a burden to maintain when in an academic research context.
3. Traditional team culture in academia, especially around team member accountability, limits the beneficial impact RPMs can have on research teams. The difference in team culture in academic versus industry settings (e.g., PIs retaining ultimate authority despite limited capacity to consistently fulfill their responsibilities) and the inability to hold all team members accountable was shared by one interview participant. “…[T]o come up with realistic suggestions for how to do this work better in an academic setting could be really potentially useful. It’s the translation from industry to academia that doesn’t quite work because of the lack of accountability” (Bethany). The differences in team member accountability structures that exist between industry environments and academia can impact the effect RPMs have on improving management practices for research projects.
4. Project management recommendations and resources that have become standard in other industries either don’t translate well to academic research environments or don’t provide the same value in academic settings. Kelli shared their experience when preparing for their Project Management Professional (PMP) certification provided by the PMI. “As I was doing the PMP, I think the PMBOK says your institution probably has templates that you can use… I was like, cool, where are our templates? I’d love to see those! And it was like, no, those actually don’t exist here…” Kevin shared how in academic settings “there wasn’t that much care for what I would call the traditional [project management] stuff.” When reflecting on their PM training and the resources they were exposed to, Lydia said “There are things out there, but they’re not quite right”, and reflected on how …“be[ing] able to tweak it so it becomes right, and relevant, and useful” would provide value. Because of the unique qualities of academic research environments, context-specific recommendations and resources are needed for managing research projects.

**Table 4:** Insights shared on the differences between academia and other industries.

| Code | Illustrative Quote | # of participants (%) |
| --- | --- | --- |
| Academic Research Prioritizes Discovery Over Product Creation | “I feel like so many of the PMI-specific resources are so tailored to software, construction, [where] you are creating a specific thing.” - *Kelli* | 2 (14.3%) |
| Academia Requires More Flexibility | “I worked previously in software development, so I'm very much focused on, like, let's create a timeline and then let's meet these deadlines and deliverables. And it seems like research requires a certain amount of flexibility for tweaking how you're going to meet the aims of your grant... It became more frustrating to try and keep this project timeline map that was historically very important for my work.” - *Kevin* | 3 (21.4%) |
| Academia has Differing Team Culture and Structure | “...[T]o come up with realistic suggestions for how to do this work better in an academic setting could be really potentially useful. It's the translation from industry to academia that doesn't quite work because of the lack of accountability.” - *Bethany* | 1 (7.1%) |
| Industry Resources Don't Translate to Academia | “There are things out there, but they're not quite right” - *Lydia* | 4 (28.6%) |

### 3.2 RPMs reported varied job responsibilities despite holding similar job titles

While most survey participants had similar job titles, this alignment was lost when it came to the description of responsibilities interview participants believe constitute project management. Sixty-two (72.9%) survey respondents shared their current job title is “project manager or equivalent title”, with 9 (10.8%) survey respondents and some interview participants sharing additional details about their specific job titles (Table 5). To ensure anonymity, full job titles are not disclosed, but the specific job titles mentioned besides “Project Manager” include terms like “Director”, “Program Manager”, “Administrator”, and “Project Specialist”.

**Table 5:** What is your job title?

| Job Title | # (%) |
| --- | --- |
| Project Manager (or equivalent title) | 62 (72.9%) |
| Other | 8 (9.4%) |
| Research staff | 8 (9.4%) |
| Research staff, Project Manager (or equivalent title) | 3 (3.5%) |
| PI or faculty member | 1 (1.2%) |
| PI or faculty member, Project Manager (or equivalent title) | 1 (1.2%) |
| Research staff, PI or faculty member, Other | 1 (1.2%) |
| Research staff, PI or faculty member, Project Manager (or equivalent title) | 1 (1.2%) |

While all survey and interview participants self-identified as RPMs or equivalent job titles, differences quickly emerged as interview participants described the specific PM activities or responsibilities they consider to be part of project management. While discussing their understanding of broader PM responsibilities, interview participants often included descriptions of their own job responsibilities, with these combined responses falling into 4 different categories of tasks: management, research, logistical, and miscellaneous tasks (Table 6).

**Table 6:** What activities or responsibilities do you consider part of project management?

| Code | Illustrative Quote | # of participants (%) |
| --- | --- | --- |
| **Management Tasks** |  | 12 (85.7%) |
| ↳ Team Members | “ We have to have very good and strong communication with the people doing the work. That is the bottom line. You have to maintain those communication channels, and you have to be respectful of what they're dealing with.” - *Ivy* | 12 (85.7%) |
| ↳ Governance | “My main thing is basically to make sure that all of our aims are delivered... I have a lot to do with the research governance.” - *Lydia* | 9 (64.3%) |
| ↳ Information | “Documenting any changes and any decisions that are made along the way, so that we have a record of how we came to where we are.” - *Nicole* | 6 (42.9%) |
| ↳ Finances | “I review all the finances.” - *Noel* | 6 (42.9%) |
| ↳ Scope | “As a project manager your role is to help filter [new project ideas] and prioritize.” - *Nora* | 4 (28.6%) |
| ↳ Physical Resources | “...[M]aking sure all the people have what they need.” - *Lacy* | 4 (28.6%) |
| ↳ Schedule | “Keeping track of scheduling.” - *Bethany* | 3 (21.4%) |
| ↳ Risk | “Making sure that I'm keeping documentation of risks.” - *Nicole* | 1 (7.1%) |
| **Research Tasks** | “I also help with the active research when we don't have enough team members” - *Lacy* | 9 (64.3%) |
| **Logistical Tasks** | “I assist investigators with both internal processes to achieve access or external processes, meaning things that, as clinicians, they may not understand a lot. Like data use agreements, business associate agreements, that there's PHI involved, or the data-related aspects of completing their IRB protocols.” - *Brandon* | 11 (78.6%) |
| **Miscellaneous Tasks** | “[I]f there's something that needs to be done and I can do it, I'll do it. Whether that's hauling a box across somewhere or something like that.” - *Drake* | 12 (85.7%) |

We coded management tasks by aligning the activity participants described with 8 PM performance domains determined by the PMI and previous research. Twelve (85.7%) interview participants shared tasks that fell into the management category which includes overseeing finances, governance, information, physical resources, risk, schedule, scope, and team members. All 12 interview participants who referenced management-related tasks cited managing team members as an activity they considered part of managing a project. One project manager reflected on this responsibility by sharing how RPMs must “have very good and strong communication with the people doing the work. That is the bottom line. You have to maintain those communication channels, and you have to be respectful of what they’re dealing with (Ivy)”. Another shared an analogy, “I really see the [project manager’s] role as sort of the air traffic control of the project. Not necessarily conducting the science, conducting analysis, conducting recruitment, etc., but it’s really… communicating to everyone who’s doing what” (Kevin).

Although some RPMs saw participating in research activities outside the scope of the role of a project manager, 9 (64.3%) interview participants shared that research-related tasks were part of an RPM’s responsibilities. These responsibilities included activities like scientific and grant writing, data collection, analysis, and management, and handling reporting requirements for funding agencies. Participants shared, “I also help with the active research when we don’t have enough team members” (Lacy) and “Some of our project managers are clinical nurses who’ve been involved with research forever, and their idea is, if the project calls for it, they’re going in there and drawing blood, or collecting samples” (Kevin).

Eleven (78.6%) interview participants cited that a project manager’s key responsibilities included logistical tasks, including administrative activities, creating project management infrastructure, and managing regulatory requirements. One interview participant shared that they “feel like a lot of project management is also administrative” and see their role as “creating some sort of structure” (Nora). Some research-specific regulatory requirements were also mentioned as key responsibilities of RPMs. “Maintaining the IRB is probably within a research project management role.” mentioned Noel, while Kevin reflected on RPMs needing to have an understanding of “data use agreements, business associate agreements, that there’s PHI involved, or more of the data-related aspects of completing their IRB protocols.” to help remove this burden from clinicians specifically.

Lastly, 12 (85.7%) interview participants mentioned that project manager responsibilities included a variety of miscellaneous tasks such as, event planning, training students, updating websites, serving as a resource connector, being a problem solver, and assistant, along with doing whatever tasks the research team needed. One project manager reflected on their role by sharing “[I]f there’s something that needs to be done and I can do it, I’ll do it. Whether that’s hauling a box across somewhere or something like that” (Drake). Drake also mentioned their role requires a lot of “being available” and “being willing to pitch in wherever”, a sentiment shared by another interview participant, Noel. Bethany described their responsibilities as “…doing anything that would fall under required work for the grant, but that’s not directly research-related. So that ends up being a lot of different things.” Some of these miscellaneous tasks included things like “organizing and running events, organizing seminars … keeping the website updated” (Bethany), and “support[ing] a lot of students and train[ing] a lot of students” (Noel). Some RPMs felt the various tasks assigned to them felt more like the job responsibilities of an assistant. “I feel like a personal assistant, right? Like, people just want to put anything that they don’t have bandwidth for, or they think is beneath them, and put it on me to do” (Nicole). As evidenced by some of these example job responsibilities, one interview participant shared “I will add that I feel like I do a lot of stuff that isn’t strictly project management” (Noel). The job title of RPM doesn’t result in clear boundaries around job responsibilities for professionals with that title.

### 3.3 Variation in job responsibilities resulted in role ambiguity

The variation in job responsibilities for RPMs highlighted by interview participants often led to role ambiguity, which was then compounded by team member unfamiliarity with working with RPMs. Together, these factors resulted in RPM’s contributions being undervalued, unclear career progression, poor utilization of the RPM’s skill set, and limited support for RPMs (Table 7).

**Table 7:** Insights shared on the challenges associated with variability in job responsibilities.

| Code | Illustrative Quote | # of participants (%) |
| --- | --- | --- |
| **Role Ambiguity** | “When I started this position, it was not very well defined. The people that hired me knew that the role needed to be filled, but they did not know what all the job was going to entail... I had to spend quite a bit of my initial time here just figuring out what was needed, because there was really not anyone that directly told me.” - *Bethany* | 6 (42.9%) |
| ↳ Team Member Unfamiliarity | “We're in the process of kicking off a quantum project and there's a lot of these documents that are required by NSF... Some of the researchers, they actually said point blank during our internal kickoff meeting, ‘Initially when I saw that NSF wanted all this, I was like, man, this is just a whole bunch of busy work... But as we're working through it, I can see the value in it, and actually having all of this documented.’” - *Kelli* | 3 (21.4%) |
| ↳ Undervalued RPM Contributions | “Nobody will pay us, but you rely on project managers to deliver the research strategy of the university. You just kind of treat us as an expensive admin.” - *Lydia* | 2 (14.3%) |
| ↳ Unclear Career Progression Pathways | “I did not know that research project management was a thing until I was well into my project management career” - *Ivy* | 5 (35.7%) |
| ↳ Poor Utilization of RPM Skill Set | “I would say as a whole about that experience for me is I feel like I am overworked, overburdened...[a]nd at the same time, I feel critically underutilized. The skill set that I have is often not maximized. The things that I have training in, I have my PMP certification, I'm a certified scrum master, all of those skills that I've worked hard to build, don't really get a chance to be taken full advantage of because I'm working on things that are at more of a project coordinator level in order to keep things moving.” - *Nicole* | 3 (21.4%) |
| ↳ Limited Support | “I am an island in some ways. I'm hungry for having a community of practice in the university context.” - *Nicole* | 4 (28.6%) |

Six (42.9%) of the RPMs interviewed shared role ambiguity as a challenge, including not only ambiguity in what the job entailed, even from as early as the initial job posting, but also misalignment between RPMs and other team members as to what the RPM’s job responsibilities are. Bethany shared how their role was unclear from the beginning. “When I started this position, it was not very well defined. The people that hired me knew that the role needed to be filled, but they did not know what all the job was going to entail… I had to spend quite a bit of my initial time here just figuring out what was needed, because there was really not anyone that directly told me.” Nicole shared their experience with misalignment with other team members about what their role on the team entails. “If I could sum it up, I wind up absorbing a consistent amount of scope creep and/or additional tasks that perhaps don’t necessarily fit in my bucket, but don’t clearly fit in any other bucket, either.” When reflecting on their experience of trying to establish clearer boundaries around what their role as a project manager entails, Nicole added “…there are some people who, when told, that’s outside of my job description, will say, ‘Okay, yep, I understand.’ And we’ll look for other ways, or look for my guidance, and who could provide that level of support. That’s the exception. More commonly, the perception is, a bit of, ‘Well, do it anyway. We need somebody to do it.’”

Three (21.4%) RPMs interviewed referenced role ambiguity stemming in part from researchers’ limited experience working with project managers for academic research projects. Nicole shared that increased confidence from PM training empowered them to establish better boundaries around their work with “a team that I felt like really didn’t have a good understanding for what a project manager actually is.” Ivy shared how they “are just kind of getting people used to working with project managers.” Role ambiguity is in part created from the limited experience of how RPMs can be utilized on research teams.

For research team members that are unfamiliar with project management practices and lack experience working with RPMs, the value of PM practices to a research project is unclear. “We’re in the process of kicking off a quantum project and there’s a lot of these documents that are required by NSF… Some of the researchers, they actually said point blank during our internal kickoff meeting, ‘Initially when I saw that NSF wanted all this, I was like, man, this is just a whole bunch of busy work… But as we’re working through it, I can see the value in it, and actually having all of this documented’”(Kelli).

The overall challenge of role ambiguity creates unclear and ill-defined pathways for career progression as an RPM. Some RPMs were unaware of what project management as a career really looked like until they were in the midst of it. “I did not know that research project management was a thing until I was well into my project management career” (Ivy). Another shared how “we didn’t know what project management was, and we’re rolling with the label” (Lacy). Irene reflected on how their job title was more of a formality connected to the work they were already doing. “[M]y official title now, I’ve had for a few months, though I’ve been doing that same job for several years and I’m just now getting a title to reflect that. And really, frankly, I’ve been doing this job for 8 or 9 years now.” Kevin shared how “it would be helpful to have some sort of enterprise standards for things. Even to the extent of defining what exactly a project manager does on projects.” They continued by highlighting the impact such a variety of job responsibilities has on the field of research project management by stating “It can be hard to write a job description that is universal… [T]herefore, if you don’t have people who have the same role with the same job responsibilities, it becomes harder to optimize what a good project manager…in research looks like.” While the lack of a defined career pathway for RPMs stems from unclear role definitions, it also further exacerbates the problem by grouping a wide variety of responsibilities under the same project manager job title.

A lack of role clarity often divided a project manager’s time and focus between competing priorities and underutilized their curated skill set, diminishing their beneficial impact on research projects. Survey and interview participants reflected on what percentage of their time is dedicated to PM activities. While 43 (50.6%) survey participants shared they spend over 75% of their time on PM activities, 8 (9.4%) indicated they spend less than 25% of their time on PM activities in a self-identified research project management role (Figure 2). Interview participants shared additional factors that influenced the percentage of time they spend on PM-related tasks, including the stage of the research project as well as other demands added to their to-do list.

**Figure 2:**
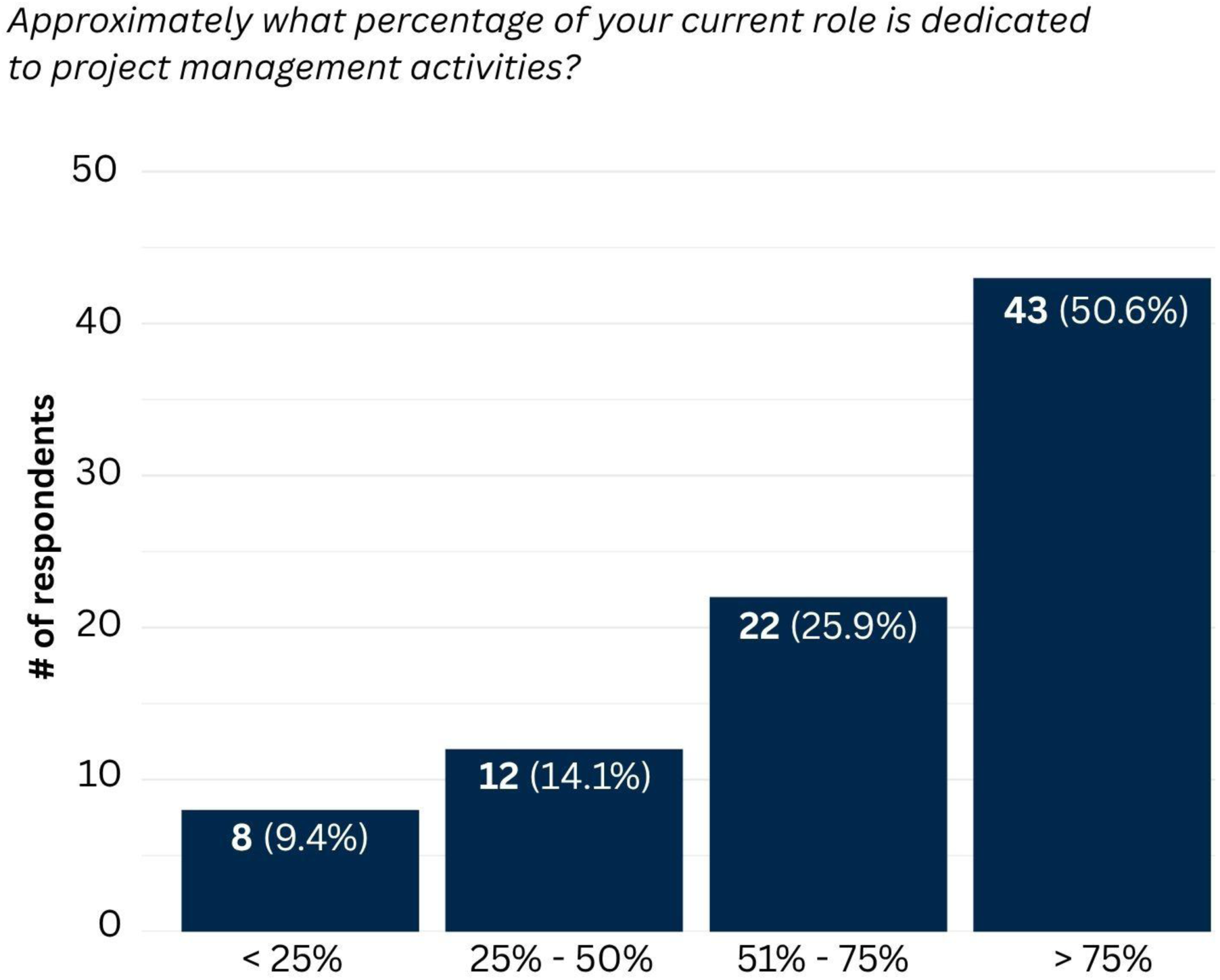
Distribution of survey respondents by percentage of time spent on project management activities in their current role. The first number indicates the number of respondents, with percentages in parentheses representing the proportion of all respondents.

Spending time and energy on tasks unrelated to project management can lead to poor utilization of RPMs on research teams. This sentiment is well summarized by Nicole who shared “I would say as a whole …I feel like I am overworked, overburdened…[a]nd at the same time, I feel critically underutilized. The skill set that I have is often not maximized. The things that I have training in, I have my PMP certification, I’m a certified scrum master, all of those skills that I’ve worked hard to build, don’t really get a chance to be taken full advantage of because I’m working on things that are at more of a project coordinator level in order to keep things moving.” This underutilization can lead to poor job satisfaction and wellbeing for RPMs. “I had to speak up and say, essentially, I’m suffering. I’m exhausted. I can’t keep up with this load.”

Lastly, 4 (28.6%) RPMs referenced how ambiguity around core job responsibilities has limited the support they have received. Some of the limitations stem from a lack of community, in which unique job titles make it difficult to identify other employees in similar roles, leading to isolation. Nicole shared “I am an island in some ways. I’m hungry for having a community of practice in the university context.” Nora added “I still love meeting other project managers. I love seeing how their brains work. It’s kind of nice because I know my boss created my role. It’s very unique…[I]it doesn’t exist anywhere. So, it’s to my benefit as well to see what other projects are going on, to stay up to date on what’s out there…” The variety in job responsibilities also impacted RPMs’ ability to further optimize resources and improve support. Kevin shared how the lack of standardization impacts the ability to share resources. “You can’t create a Research Project Management Resource Page if it’s not clear every RPM hears the list of things they would need.” Nicole shared how due to the expansion of their core responsibilities and being overworked, they “…pushed and pushed for the past year that I’ve been on this project for a coordinator…”, and it has been difficult getting support to hire the help necessary. RPMs in academic settings face unique challenges and want support to improve not only their own well-being and job satisfaction, but the beneficial impact they have on CTR projects.

### 3.4 RPMs have identified key areas of support that can improve how research projects are managed

Through reflections on their experience as RPMs in academic environments, 12 (85.7%) interview participants identified support and specific resources they believe can improve how research projects are managed. These insights were organized from broad, system-level support to more individual role-specific recommendations (Figure 3) and include: (1) improved accountability structures within academic research, (2) dedicated funding allocation to support project management systems, (3) updated hiring practices, (4) project management onboarding and training, (5) access to useful project management software and integrations, (6) peer support through communities of practice, and (7) project management recommendations and resources tailored to an academic research environment (Table 8).

**Figure 3:**
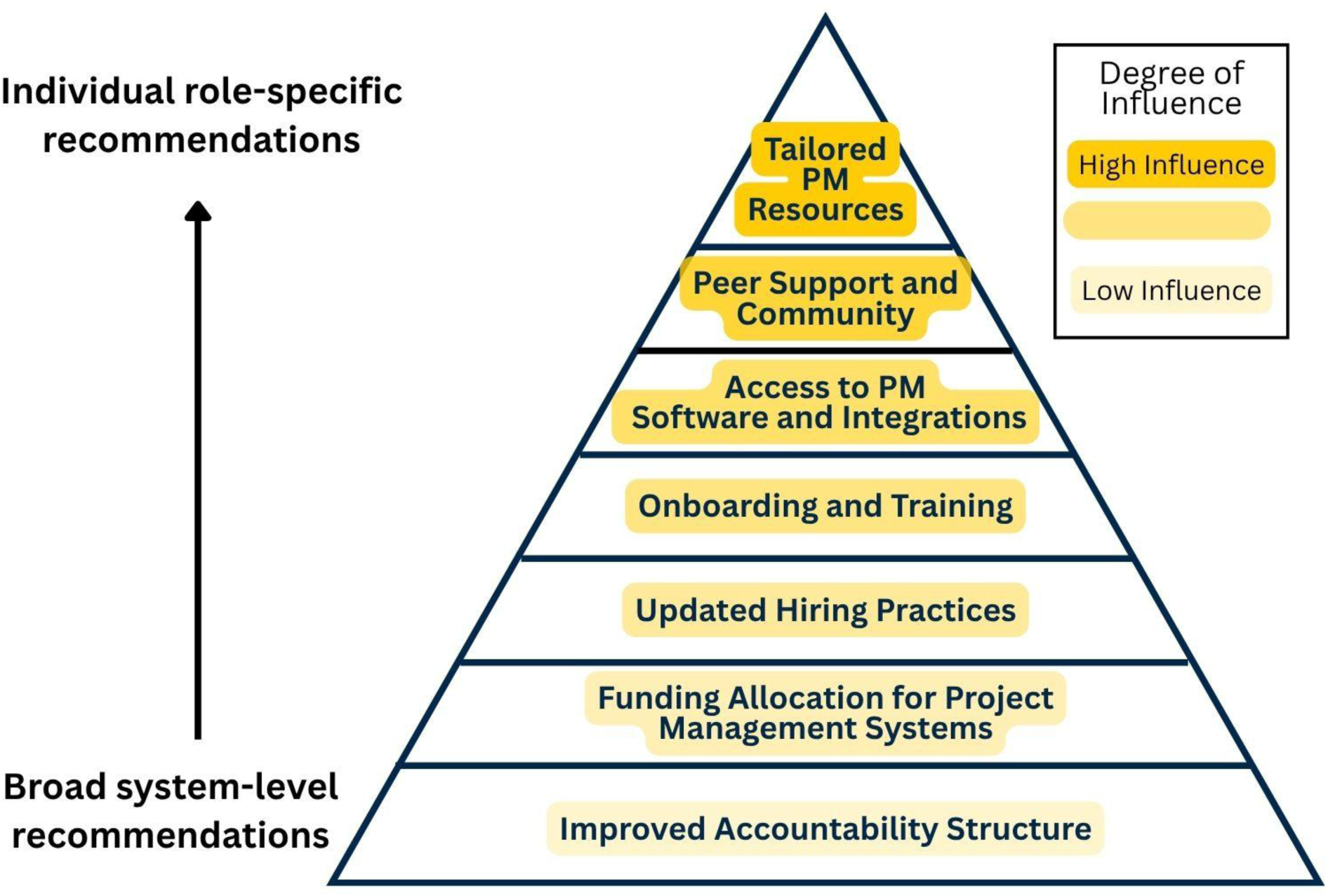
Hierarchy of recommendations to improve research project management shared by interview participants. Recommendations were arranged from broad, system-level suggestions at the base of the triangle to individual, role-specific suggestions at the top. The color gradient represents the relative degree of influence an individual may have over each recommendation, with lighter colors indicating less influence and darker colors indicating greater influence.

**Table 8:** What resources do you think would be useful to improve project management for research projects like the one you are currently working on?

| Code | Illustrative Quote | # of participants (%) |
| --- | --- | --- |
| Improved Accountability Structure | “Unless you can really change the accountability structure for faculty on these grants, I don't know how you can really produce a lot of buy-in from them to do things the way that you as a project manager or as a funding agency would like them to be done.” - *Bethany* | 1 (7.1%) |
| Funding Allocation for PM Systems | “[Funding agencies] making the recommendation that some amount of your budget goes towards not just the salary of the project manager, but to the structures and services that will help the project manager to lead the team.” - *Nicole* | 1 (7.1%) |
| Updated Hiring Practices | “[H]aving a dedicated project manager. It seems like a lot of research projects don't have a dedicated project PM, they just have the PI that maybe doesn't have the necessary training. And when you get into the bigger [projects], especially cooperative agreements that have broadening participation requirements and outreach and workforce development and things like that, it's a lot to manage. So I think if you have someone that's solely dedicated to that administrative piece, it'll help a lot.” - *Brandon* | 2 (14.3%) |
| Onboarding and Training | ““[H]aving a system only works if people know how to use it, so having the training to go along with it. And the training not just for people using it, but for the people they report to.” - *Irene* | 4 (28.6%) |
| ↳ Greater Resource Awareness | “I guess I'm not entirely sure about what would be out there that could help me.” - *Drake* | 3 (21.4%) |
| Access to PM Software & Integrations | “When I was at [my previous institution], we were able to have a Google Drive integration in Box, and that was the best of all worlds for me. And when I moved here, I tried | 2 (14.3%) |
|  | so hard to see if that was something we could get done here. Our IT team won't do it for security reasons, which is understandable, but... it made everything possible to be in one system..." - *Nora* |  |
| Peer Support and Community | "[A]... learning circle and have other contacts at the university that [project managers] can network with which is great for employee well-being" - *Nora* | 6 (42.9%) |
| Tailored Recommendations and Strategies | "A toolbox, or something for project management, specifically for research projects, I think would be so helpful." - *Lacy* | 5 (35.7%) |
| ↳ Standardized Templates and Resources | "Ideally, from supporting research, I would say it would be helpful to have some sort of enterprise standards for things." - *Kevin* | 2 (14.3%) |

Interview participants identified broad support that would improve project management for research projects. Bethany, who previously shared how the structure of accountability on research teams contributes to the differences between academia and other industries, expressed their desire to see a change in the accountability structure within academic institutions. “What it comes down to is, unless you can really change the accountability structure for faculty on these grants, I don’t know how you can really produce a lot of buy-in from them to do things the way that you as a project manager or as a funding agency would like them to be done.” In addition to seeing a broader shift in some of the team structures that exist within academia, Nicole highlighted one way funding agencies specifically could support improved PM practices by “making the recommendation that some amount of [the proposed] budget goes towards not just the salary of the project manager, but [to] the structures and services that will help the project manager to lead the team”.

Some interview participants shared intermediate-level support suggestions that could have a beneficial impact on academic research projects. Brandon shared the suggestion of “having a dedicated project manager. It seems like a lot of research projects don’t have a dedicated project PM; they just have the PI that maybe doesn’t have the necessary training. And when you get into the bigger [projects], especially cooperative agreements that have broadening participation requirements and outreach and workforce development and things like that, it’s a lot to manage. So, I think if you have someone that’s solely dedicated to that administrative piece, it’ll help a lot.” Ivy added that more PIs should “Write [project managers] into your grants. Write us into your proposals.” Additional impact to CTR would be created through ensuring RPMs are able to record project knowledge and lessons learned even after grant funding has ended. “The magic wand would be to have some kind of financial support structure that allows [research project managers] to do all of the post-closeout activities, like the post-mortems, like creating reusable artifacts for future projects” (Ivy). Updating hiring practices to include RPMs on more projects and ensure they have financial support outside of a grant can have a beneficial impact on how research projects are managed and how the lessons learned from previous projects are preserved.

Some RPMs discussed providing onboarding for newly hired project managers as well as PM training not only for RPMs, but their team members as well. Nora shared that what they would find useful “would be for institutions to create [project management] training courses for new hires.” Irene shared how these trainings would connect to PM systems by saying, “[H]aving a system only works if people know how to use it, so having the training to go along with it. And the training not just for people using it, but for the people they report to.” Onboarding and training would not only be beneficial for sharing more about project management and the value it can provide to a research team, but also exposing new hires and researchers to the tools and resources around project management that exist at their institutions. When answering the question about what resources RPMs thought would be useful to improve project management for research projects, some struggled to provide an answer because they weren’t “entirely sure about what would be out there that could help me” (Drake). Irene shared their experience with learning about resources available at their institution, “We connect people to resources that exist here at [my institution] [that] they don’t know about. I’m still learning about resources. I’ve been here for 17 years. There’s still many resources I do not know exist. And I only learned because friends of mine are like, ‘Have you seen this?’”. Improving onboarding practices and increasing PM training for all members of academic research teams impacts research projects and exposes all team members to available tools and resources that can enhance their work.

RPMs we interviewed also mentioned how institutions could support the management of research projects by providing access to software and allowing integrations between different software companies. One project manager shared how switching institutions required losing some of the positive functionality in a previously established workflow. “When I was at [my previous institution], we were able to have a Google Drive integration in Box, and that was the best of all worlds for me. And when I moved here, I tried so hard to see if that was something we could get done here. Our IT team won’t do it for security reasons, which is understandable, but… it made everything possible to be in one system…” (Nora). Another interview participant shared “my university definitely will not entertain, covering the cost for licenses for things like that that a project manager might want.” By ensuring research teams have access to a variety of software and integrations commonly employed by RPMs, institutions can help streamline research project workflows.

Lastly, interview participants shared support suggestions that would benefit their individual roles as RPMs in both job satisfaction and professional development, including the value of increased community amongst other project managers in academic research environments. Nora shared that it would be beneficial to have a “…learning circle and have other contacts at the university that [project managers] can network with which is great for employee well-being”. Brandon shared “being able to look into projects that do have a PM and see what they’ve done differently from your project that doesn’t have a PM might be helpful.” Bethany shared that going beyond community and having regular guidance from a mentor would be valuable. “Mentorship from people who have had similar roles in the past is quite useful.” In addition to having increased community with other RPMs, interview participants highlighted the desire to have standardized recommendations, strategies, and resources tailored to research contexts. Kevin shared, “Ideally, from supporting research, I would say it would be helpful to have some sort of enterprise standards for things.” Lacy added “A toolbox, or something for project management, specifically for research projects, I think would be so helpful.”

To understand the potential value of PM recommendations and resources tailored to an academic research environment, we asked survey participants to share on a scale of 1–5, how likely they would be to use a project management resource or tool tailored to an academic research context, which resulted in a mean score of 3.94 (standard deviation 1.08). Interview participants added in context about the usefulness of resources tailored to academia. While one interview participant shared, “I don’t really think [resources] would have to be specifically tailored to the research environment” (Brandon), others highlighted benefits of improved efficiency through saved time and increased focus, better research and work environment, easier onboarding processes for new hires, and improved research outcomes through stronger foundational knowledge (Table 9). One participant shared, “I feel like it makes better environments for people… If you’re always putting fires out and you’re just winging it, that doesn’t make for a fun environment for staff or for the researcher who wants to be doing research, not trying to manage bureaucracy within the institution, or figuring out how to get your sponsored projects office to pay an invoice. Some of that stuff, it’s just a drag to the actual work of getting research innovations and discoveries out. It’s about efficiency, it’s about creating good environments that are productive and fulfilling careers, and, for staff and faculty, being able to do a job well” (Lucy). Receiving support can not only improve the well-being and job satisfaction of RPMs but can have broader impacts for the entire research team and the clinical and health outcomes their research contributes to.

**Table 9:** What are some benefits you can imagine of having project management resources that are better tailored to an academic research environment?

| Listed Benefits | Illustrative Quote | # of participants (%) |
| --- | --- | --- |
| Saves Time | “If everything was a little bit more organized and a little bit more streamlined, I would have more time to do the things I needed to do.” - *Lacy* | 5 (35.7%) |
| Better Work Environment and Job Satisfaction | “I feel like it makes better environments for people... If you're always putting fires out and you're just winging it, that doesn't make for a fun environment for staff or for the researcher who wants to be doing research, not trying to manage bureaucracy within the institution, or figuring out how to get your sponsored projects office to pay an invoice. Some of that stuff, it's just a drag to the actual work of getting research innovations and discoveries out. It's about efficiency, it's about creating good environments that are productive and fulfilling careers, and, for staff and faculty, being able to do a job well” - *Lucy* | 4 (28.6%) |
| Improves Efficiency | “I feel like there is responsibility to taxpayers to be efficient in doing good project management so that the money is being used for discovery and not for redundancies and inefficiencies.” - *Lucy* | 3 (21.4%) |
| Easier Onboarding Process | “I think as a new hire, it would have been, like, priceless.” - *Nora* | 3 (21.4%) |
| Improves Research | “It makes science so much better. It keeps us all focused.” - *Ivy* | 1 (7.1%) |
| Increases Focus | “It starts with the PIs, convincing them that a project manager is going to allow you to do a lot more work, and you don't have to be in the weeds sending out the emails or sending the reports.” - *Ivy* | 1 (7.1%) |
| Builds Knowledge Base | “[Y]ou also then start to build a knowledge base...If you're documenting what's been learned already, that's just more resource there to feed [a personal project manager chatbot]. Quicker answers, quicker solutions, moving on to the next aspect of a research project.” - *Kevin* | 1 (7.1%) |
| Tailoring<br>Resources is<br>Unnecessary | “I don't really think [resources] would have to be specifically tailored to the research environment.” - *Brandon* | 1 (7.1%) |

## 4 Discussion

Our analysis of survey and interview data from RPMs in academic environments yielded four main results: 1) RPMs showed alignment in their definition of project management and highlighted key differences between project management in academia versus other industries. 2) Although there were high degrees of similarity in job titles, there was a wide variety of specific job responsibilities represented. 3) Variation in job responsibilities between RPMs resulted in struggles with role ambiguity. 4) Participants in this study highlighted key areas of support that can improve how academic research projects are managed.

The beneficial impact good PM practices can have on a research project are well documented, however the role of RPMs in academia still lacks clarity (Pacheco Junior et al., 2020; Payne et al., 2011). Funding agencies have begun acknowledging the importance of well-managed projects in achieving research goals. This is evident in the requirement of a management plan for some grant proposals like the NSF’s Science and Technology Centers: Integrative Partnerships (U.S. National Science Foundation, 2024). This requirement would be strengthened, however, by providing a clearer description of what job responsibilities the project manager for these proposed research projects should have. Our study highlighted the wide variety of tasks and roles RPMs can play in academia and the unique challenges that come from this breadth. While some variety is necessary and inevitable, CTR teams will see the greatest impact of RPMs when there is clarity around their job responsibilities and clearer boundaries between different roles to ensure the skill set RPMs provide is utilized well. Some research groups have already proposed potential career tracks for RPMs to help establish these role boundaries, however, more work is needed to further solidify this career path and more fundamentally define the roles and responsibilities of RPMs (Sutton et al., 2019; Sweeney et al., 2025).

While defining research project management, acknowledging the value of good PM practices on research outcomes, and standardizing the roles and responsibilities within research project management as a career are important initial steps, RPMs need additional support to maximize their beneficial impact on research projects. Our study highlighted some of these suggestions from RPMs themselves (Table 8). We organized these suggestions from broad, system-level recommendations to more individual, role-specific suggestions, however, the interconnectedness of funding agencies, institutions, and research teams makes it difficult to know where to start or who should be responsible for leading effectual change toward any of these suggestions. While organizing suggestions for how to improve research project management, it became clear that suggestions that were most individual and role-specific were the ones where an individual would have the highest amount of influence to effect change (Figure 3). The two recommendations that serve as a first point of intervention for supporting RPMs are those with the highest level of individual influence: peer support and community and tailored PM resources.

Results from our study highlighted the value that community can have for RPMs. Although there are some communities like the International Network of Research Project Managers (INRPM) and National Research Project Management Network (RPM-Net) emerging more broadly as support networks for those in research project management positions, having peer support from those working within the same institution can provide a more context-specific level of assistance and mentorship. Institutions like Duke University have created an established PM community of practice within their institution that has large engagement of members and serves as a good example of the power of community and the desire for employees to learn and grow from peers within their institution (Johnson et al., 2020). Providing avenues for connection and community amongst RPMs is an area where involvement from invested individuals can lead to quick progress and incorporation of this suggestion.

Another suggestion with high individual influence is creation and broad dissemination of PM resources, strategies, and recommendations that are tailored to an academic research context. While organizations like the PMI provide a wide variety of resources to improve project management, some of the unique qualities of academia highlighted in this study make those recommendations difficult for RPMs to easily employ. While creation of tailored resources is a great point of intervention for supporting RPMs, it requires additional research to decide which resources should be tailored to an academic research context, determine how to tailor resources for maximal effect, and assess the impact these resources have on research outcomes.

### 4.1 Future Directions

This study revealed areas for future study around research project management and points of intervention where individuals have the highest level of influence. Future studies and input are required from the entire research community, including not only RPMs, but PIs, administrators, funding agencies, and institutions to address some of this study’s key findings including: a definition of research project management agreed upon by consensus; a catalog of standardized roles, responsibilities, and career progression pathways for RPMs; and creation of support networks and community for those in this profession. This standardization could greatly improve the work environment, job satisfaction, and influence RPMs have on progressing collaborative research projects from ideas to practice. Future studies should explore the impact these suggestions have not only on the field of research project management, but on research environments and outcomes more broadly. As noted above, creating PM resources and recommendations tailored to an academic research context would be a valuable point of intervention to help improve the impact and efficiency of CTR projects.

Overall, what this study highlights is the unique role RPMs have, and the lack of resources designed to support this professional group directly and explicitly. Because of the unique qualities of academic research, future publications of the dataset used for this manuscript will aim to identify the challenges RPMs specifically face when managing research projects and gain a better understanding of the tools and techniques RPMs currently employ. After identifying the main challenges RPMs face within academia, we will identify resources and recommendations from other industries to address these issues and engage the research project management community to determine what adjustments might be necessary to better fit these resources to an academic research environment. Lastly, we not only want to provide more tailored support and guidance to RPMs on ways to improve management for their research projects but also demonstrate the impact this tailored guidance has on research productivity and team member wellbeing. Future studies will aim to create tailored resources and training around project management for CTR teams and test the efficacy of implementing these resources by assessing metrics like publication count, grant funding received, perceived research productivity, job satisfaction, and team member psychological safety.

### 4.2 Conclusion

As CTR increases in complexity, successful translation of discoveries into interventions will require effective management of research projects. When good management practices are applied to a project, their impact is often invisible: steady progress is made on work related tasks, team members are clear on their role in the project, roadblocks and issues are quickly addressed, and the project moves forward smoothly. Well managed projects rarely happen naturally. There is normally someone driving the process of turning chaos into order. For many CTR projects, especially those with large, interdisciplinary teams, this process of helping a project progress smoothly is often led by RPMs. The often intangible impact RPMs provide to the completion of research projects can leave the people in these roles feeling invisible. Our study shined a light on RPMs and highlighted the ambiguity that currently exists around the role and responsibilities of RPMs. While this lack of clarity creates unique challenges for RPMs, they have suggestions for necessary support they would like to receive to maximize their impact on CTR projects. By focusing on RPMs, we not only highlight the important role these team members play in progressing important research initiatives, we also identify ways to improve workplace environments for every member of a research team and the impact of research outcomes. The smoother research projects run and the better supported all team members feel, the faster impactful research gets translated from the bench to the bedside.

## Data Availability

All data produced in the present study are available upon reasonable request to the authors.

## 5 Data Availability Statement

The datasets for this study are not currently available as this is the first in a series of papers and the codebook is still under development. Once finalized, the codebook will be made available in future manuscripts. Requests to access the dataset should be directed to Shayla Vradenburgh.

## 7 Author Contributions

SAV: Conceptualization, Data curation, Formal analysis, Investigation, Methodology, Project administration, Visualization, Writing - original draft.

BR: Conceptualization, Funding acquisition, Methodology, Supervision, Visualization, Writing - review & editing.

## 9 Generative AI Statement

The author(s) declared that generative AI was used in the creation of this manuscript. The author(s) used OpenAI’s ChatGPT to assist with word choice and text editing for original writing and in creating clear and concise code labels for more complex themes.

## 10 Acknowledgements

We would like to thank Shruthi Venkatesh for support and feedback on this manuscript. We thank Liz Fisher for recruitment support, and Elizabeth LaPensee for general project oversight.

