## Supplementary Material for "Fragmented and Underutilized: Exploring Role Variation and Ambiguity for Research Project Managers in Collaborative Team Science"

### **1 Survey**

1. What is your job title? (Select all that apply)
   a. Undergraduate Student
   b. Graduate student
   c. Postdoc
   d. Research staff
   e. PI or faculty member
   f. Project Manager (or equivalent title)
   g. Other
2. Approximately how long have you been in your current position?
   a. Less than 1 year
   b. 1 to 2 years
   c. 3 to 5 years
   d. 6 to 10 years
   e. More than 10 years
3. What best describes the institutional home of your primary project?
   a. US-based research-focused institution (R1)
   b. Other US-based university or college
   c. Non-US university or college
   d. Independent research institution
   e. For-profit research institution
   f. Government research organization
   g. Other
4. What kind of grant funding supports your primary project? (Select all that apply)
   a. Federal
   b. Foundation
   c. Institutional
   d. Industry-sponsored
   e. No external funding
   f. Other
5. What is your primary research area?
   a. Biomedical Science
   b. Behavioral and Social Science
   c. Clinical Science
   d. Other
6. What type(s) of research does your primary project involve? (Select all that apply)
   a. Basic
   b. Clinical
   c. Translational
   d. Community-engaged
   e. Other
7. Approximately how many people are involved in your primary project team (including yourself)?
   a. 1 (just me)
   b. 2 to 5
   c. 6 to 10
   d. 11 to 20
   e. More than 20
8. How are team members distributed across labs or organizations for your primary project?
   a. Primarily within one lab
   b. Across multiple labs within the same institution
   c. Across multiple labs at different institutions
   d. Collaborative project with an external industry or company partner
   e. Other
9. Approximately how many other projects are you involved in where your role is similar to your role on your primary project?
   a. 1
   b. 2 to 3
   c. 4 to 6
   d. 7 to 10
   e. More than 10
10. Approximately how many projects are you involved in overall (including projects where your role differs from your primary project)?
    a. 1
    b. 2 to 3
    c. 4 to 6
    d. 7 to 10
    e. More than 10
11. The Project Management Institute defines project management as “the application of knowledge, skills, tools, and techniques to project activities to meet project requirements. ”On a scale from 1 to 5, rate how well this definition is aligned with your experience and understanding of project management.
    a. 1 (No Alignment)
    b. 2
    c. 3 (Moderately Aligned)
    d. 4
    e. 5 (Extremely Aligned)
12. How would you describe your role on your primary project?
    a. Project manager (or equivalent role)
    b. Primary researcher (driving day-to-day scientific work)
    c. Both project manager and primary researcher
    d. Other
13. Have you ever received any formal training in project management?
    a. Yes
    b. No

13A. On a scale of 1 to 5, how impactful has formal project management training been on your research productivity?

a. 1 (Not Impactful)
b. 2
c. 3 (Moderately Impactful)
d. 4
e. 5 (Extremely Impactful)

13B. On a scale of 1 to 5, how impactful do you believe formal project management training would be for your research productivity?

a. 1 (Not Impactful)
b. 2
c. 3 (Moderately Impactful)
d. 4
e. 5 (Extremely Impactful)
14. In a few words, what is the biggest project management challenge you have encountered on your primary project?
15. For your primary project, please rank the following project management responsibilities from most challenging (1) to least challenging (10):
    a. Managing Team Members (communicating expectations and responsibilities and discussing progress)
    b. Managing Finances (budgeting, tracking expenses, and allocating funds)
    c. Managing Scope (defining project goals, deliverables, and boundaries)
    d. Managing Collaborators (coordinating contributors’ roles and aligning skills with project needs)
    e. Managing Physical Resources (procuring and maintaining supplies, equipment, and materials)
    f. Managing Time (scheduling and executing tasks and ensuring deadlines are met)
    g. Managing Quality (ensuring work products meet established standards and requirements)
    h. Managing Information and Data (creating, storing, organizing, and accessing project data and documentation)
    i. Managing Internal Risk Factors (identifying and addressing barriers within a lab or institution such as policies, team dynamics, or institutional constraints)
    j. Managing External Risk Factors (monitoring and responding to influences external to your institution such as federal funding changes or regulatory shifts)
16. Based on the project management responsibilities in the previous question, approximately what percentage of your current role is dedicated to project management activities?
    a. < 25%
    b. 25%–50%
    c. 51%–75%
    d. > 75%
17. Which project management tools or resources do you regularly use for your primary project? (Select all that apply)
    a. No tools used
    b. Analog tools (e.g., pen and paper, whiteboard, printed binders)
    c. Spreadsheets (e.g., Excel, Google Sheets)
    d. Email
    e. Shared file storage (e.g., Google Drive, Box, OneDrive)
    f. Team chat (e.g., Slack, Teams)
    g. Project management platform (e.g., Asana, Trello, Monday, Notion)
    h. Calendar/scheduling tools
18. Thinking about the project management resources or tools you currently use (if any), how would you describe your experience?
    a. I use project management tools or resources regularly.
    b. I find the tools or resources I use easy to navigate.
    c. I find project management tools or resources unnecessarily complex.
    d. I find project management tools or resources cumbersome to use in practice.
    e. I feel confident in my ability to use project management tools or resources effectively.
    f. I typically need support or guidance to use project management tools or resources.
    g. I feel that project management tools or resources improve how I manage my work.
    h. I do not currently use any formal project management tools or resources.
19. Which statement best describes how you access or create project management resources or tools?
    a. I primarily use tools that are provided by my lab, program, or institution.
    b. I primarily pay for my own tools (personally or using discretionary funds).
    c. I primarily create or build my own tools (e.g., spreadsheets, trackers, templates).
    d. I do not use any tools.
20. Where did you hear about the resources or tools that have been most impactful in your research productivity?
    a. Personal research
    b. Colleagues or friends
    c. Project management seminars/workshops
    d. Formal project management organizations (e.g. PMI, IPMA, Axelos)
    e. Other
21. For your primary research project, how do you most often use project management tools to manage work?
    a. I use these tools primarily to manage my own work (I am not currently working with a team, or I rarely use these tools with others).
    b. I am the primary person who enters and updates information for the team (e.g., tasks, timelines, assignments).
    c. Information entry and updates are shared across the team (multiple team members regularly add/update items).
    d. I primarily view or reference information entered by others (I rarely add or update items myself).
    e. I do not use any project management tools.
    f. Other
22. Approximately what percentage of your primary project team uses the same project management tool(s) you use?
    a. 0% (none)
    b. 1%–25%
    c. 25%–50%
    d. 51%–75%
    e. > 75%
23. On a scale of 1–5, how likely would you be to use a project management resource or tool tailored to an academic research context?
    a. 1 (Not At All Likely)
    b. 2
    c. 3
    d. 4
    e. 5 (Extremely Likely)
24. If a project management resource or tool were tailored to your specific research environment, how do you think you would experience using it? (Select all that apply)
    a. I would use a tailored project management resource frequently.
    b. I would expect it to be easy to use.
    c. I would expect it to be difficult to learn or navigate.
    d. I would need support or training to use it effectively.
    e. I would need a better understanding of project management concepts before using it.
    f. I would feel confident using it on my own.
    g. I would expect it to streamline or simplify my work.
    h. I would find it cumbersome to integrate into my workflow.
    i. My current project management workflow works well for me; I do not feel a need for additional or tailored resources.
